# Combined translational and rotational perturbations of standing balance reveal contributions of reduced reciprocal inhibition to balance impairments in children with cerebral palsy

**DOI:** 10.1101/2023.03.28.23287868

**Authors:** Willaert Jente, Desloovere Kaat, Van Campenhout Anja, Lena H. Ting, De Groote Friedl

## Abstract

Balance impairments are common in cerebral palsy (CP). When balance is perturbed by backward support surface translations, children with CP have increased co-activation of the plantar flexors and tibialis anterior muscle as compared to typically developing (TD) children. However, it is unclear whether increased muscle co-activation is used as a compensation strategy to improve balance control or is a consequence of impaired reciprocal inhibition. During translational perturbations, increased joint stiffness due to co-activation might aid standing balance control by resisting movement of the body with respect to the feet. However, during rotational perturbations, increased joint stiffness will hinder balance control as it couples body to platform rotation. Hence, we expect increased muscle co-activation in response to rotational perturbations if co-activation is caused by reduced reciprocal inhibition but not if it is merely a compensation strategy.

We perturbed standing balance by combined backward translational and toe-up rotational perturbations in 20 children with CP and 20 TD children. Our perturbation protocol induced a backward movement of the center of mass requiring balance correcting activity in the plantar flexors followed by a forward movement of the center of mass requiring balance correcting activity in the tibialis anterior.

We found that the switch from plantar flexor to tibialis anterior activity upon reversal of the center of mass movement was less pronounced in children with CP than in TD children leading to increased co-activation of the plantar flexors and tibialis anterior throughout the response. Therefore, our results suggest that a reduction in reciprocal inhibition causes muscle co-activation in reactive standing balance in CP.

## INTRODUCTION

Balance impairments are common in cerebral palsy (CP), but it is yet unclear which motor control pathways contribute to these balance impairments (1,2). When balance is perturbed by backward support surface translations, children with cerebral palsy have increased co-activation of the plantar flexors and tibialis anterior muscle as compared to typically developing (TD) children (1–4). However, it is unclear whether muscle co-activation is a compensation strategy to improve balance control or a consequence of impaired reciprocal inhibition (5,6). Increased joint stiffness due to muscle co-activation might aid standing balance control in response to translational perturbations by resisting movement of the body with respect to the feet. However, increased joint stiffness will hinder standing balance control in response to rotational perturbations as it induces coupling between the body and platform motion resulting in body tilt. Hence, if co-activation is used as a compensation strategy in children with CP in translational perturbations, we do not expect to observe it in response to rotational perturbations. But if co-activation is caused by impaired reciprocal inhibition, we do expect to observe it in response to rotational perturbations as well.

Reduced reciprocal inhibition is often observed in cerebral palsy (6,7). Reciprocal inhibition is defined as the inhibition of antagonistic muscle activity upon agonistic muscle activation (8). In healthy adults, rapid dorsiflexion of the ankle (i.e., stretching the plantar flexors) induces a strong response in the stretched muscle, while the antagonistic muscle (i.e., tibialis anterior) remains silent. In children with CP, such strong response is also observed in the antagonistic muscle and the responses in agonistic and antagonistic muscles have similar latencies. This impaired reciprocal inhibition in CP is observed both at rest and with different levels of muscle activation (5,6). A reduction in reciprocal inhibition might explain altered sensorimotor processing in cerebral palsy, i.e., alterations in how the nervous system translates incoming sensory information about body motion into motor commands to activate muscles.

We recently found that muscle activity in response to backward support surface translations during standing can be explained by delayed feedback of center of mass (CoM) kinematics in both TD children and children with CP, yet both agonists and antagonists were more sensitive to CoM disturbances in children with CP (4). Inspired by previous work in healthy and pathological humans and animals (9–13), we reconstructed reactive muscle activity of the plantar flexors by a weighted linear combination of delayed (to account for neural transmission times) CoM displacement, velocity, and acceleration. For the tibialis anterior, we included both a stabilizing pathway that activates the muscle when the CoM moves backward and a destabilizing pathway that activated the muscle when the CoM moves forward (12). The weights or gains of the CoM displacement, velocity, and acceleration in the linear combination indicate the sensitivity of the muscle response to CoM disturbances. We found that displacement and velocity gains were higher for plantar flexors and tibialis anterior in children with CP than in TD children (4). The higher gains for the tibialis anterior for the destabilizing pathway reflect increased co-activation and might reflect reduced reciprocal inhibition. Yet, to distinguish reduced reciprocal inhibition from the use of co-activation as a successful compensation strategy, we should also investigate the response to rotational perturbations since muscle co-activation will not help balance recovery during rotational perturbations.

In healthy humans and animals, CoM movement seems to dictate muscle responses to a wide range of translational and rotational support-surface perturbations. Translational and rotational perturbations that elicit similar CoM movements yet different joint movements induce activity in similar groups of muscles (13–16). Furthermore, Safavynia & Ting (10) found that delayed feedback from CoM kinematics but not joint angles could explain reactive muscle activity during a train of translational perturbations that induced uncorrelated CoM and joint angle trajectories. By combining backward translational and toe-up rotational perturbations, we can induce a change in the direction of the CoM displacement during the response (forward in response to translation followed by backward in response to rotation). Based on the previously proposed CoM feedback theory, a switch from plantar flexor to tibialis anterior muscle activity upon reversal of the CoM displacement is expected in healthy subjects. However, if reciprocal inhibition is impaired in children with CP, we expect them to have trouble with switching from plantar flexor to tibialis anterior muscle activity upon reversal of the CoM displacement.

Here, we combined translational and rotational perturbations to test whether reduced reciprocal inhibition can explain alterations in balance control in children with cerebral palsy. If reciprocal inhibition is reduced in children with CP, we expect to see increased co-activation and more specifically a reduced ability to suppress plantar flexor activity upon reversal of the CoM movement in response to combined backward translational and toe up rotational perturbations. First, we evaluated reactive muscle activity. We hypothesized that children with CP would have prolonged activity in the plantar flexors upon reversal of the CoM movement as well as increased muscle co-activation. Second, we evaluated the relation between CoM movement and reactive muscle activity. The sensorimotor model used for this analysis describes both a stabilizing and destabilizing pathway based on CoM movement and is therefore suitable to distinguish a muscle’s role as an agonist and antagonist. We hypothesized that CoM feedback could explain muscle activity in both TD children and children with CP but that gains for the stabilizing and destabilizing pathway would be higher in children with cerebral palsy.

## METHODS

### Subjects

The study was approved by the Ethical Committee of UZ/KU Leuven (S63321). Forty-six children participated in this study. A legal representative of the participant and participants signed respectively an informed consent or informed assent form before the start of the measurements according to the principles of the Declaration of Helsinki. Children with CP were recruited through the CP reference center at the University Hospital Leuven (Belgium). All patients were diagnosed as having spastic CP by a neuro-pediatrician and met the following inclusion criteria: (1) 5 to 17 years old; (2) Gross Motor Function Classification Scale (GMFCS) I-III; (3) able to stand independently for at least 10 minutes; (4) no orthopedic or neurological surgery in the previous year; and (5) no Botulinum Toxin injections in the previous 6 months. TD children were recruited through colleagues and friends and were age matched with the children with CP.

Data from six children were excluded due to (1) the child not completing the whole protocol (N = 1), (2) the child being unable to follow the instructions (N = 3), or (3) missing EMG data due to technical errors (N = 2). Data from 20 TD children (8 girls/12 boys) and 20 children with CP (9 girls/11 boys) were included for further analysis (table 1). Fourteen children with CP were unilaterally involved and six children with CP were bilaterally involved. Fifteen children had GMFCS level I and five children had GMFCS level II. Sixteen children had spasticity in the gastrocnemius muscle, as indicated by a Modified Ashworth Scale between 1 and 3 (more information in supplementary material S1 (table S1)). All children were able to stand and walk without walking aids. For the children with CP, the most affected leg based on clinical spasticity scores was used for further analysis, while for TD children one leg was randomly selected.

**Table 1:**
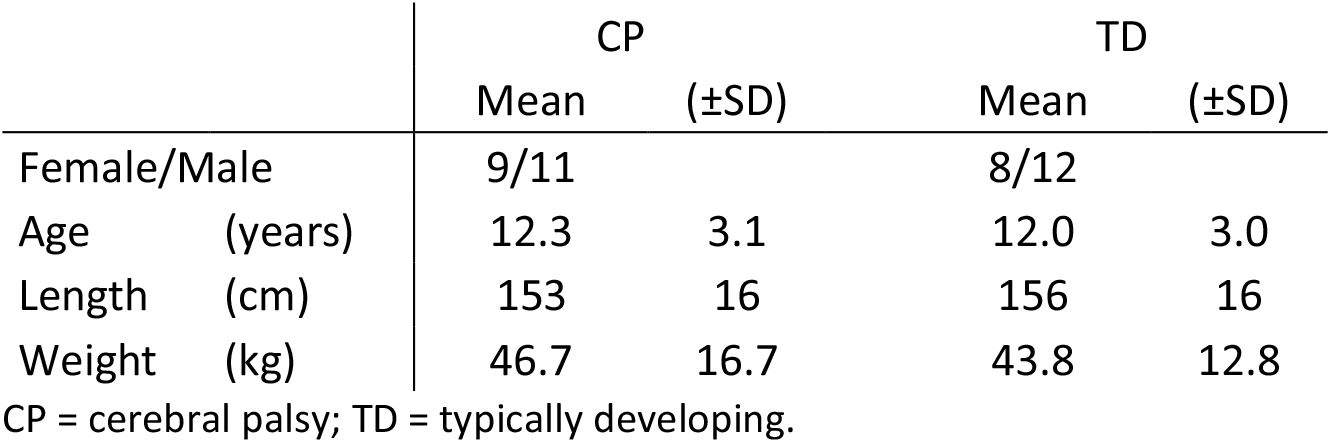
Demographic data of participants (mean and standard deviations)

### Materials

Trajectories of reflective skin markers (for details on marker placement, see supplementary material S2 (figure S1)) were captured by 7 infrared Vicon cameras (Vicon, Oxford Metrics, United Kingdom, 100 Hz). Activity of gastrocnemius lateralis (LG), gastrocnemius medialis (MG), soleus (SOL), and tibialis anterior (TA) was measured simultaneously through surface electromyography (sEMG, ZeroWire EMG Aurion, Cometa, Italy, 1000 Hz). Silver-chloride, pre-gelled bipolar electrodes (Ambu Blue Sensor, Ballerup, Denmark) were placed according to SENIAM guidelines (17). Reactive balance was tested using combined translational and rotational perturbations on an instrumented, movable platform (Caren platform, Motek, The Netherlands) (figure 1a). Children were secured using a safety harness, adjusted to an overhead rail to prevent falling.

**Figure 1:**
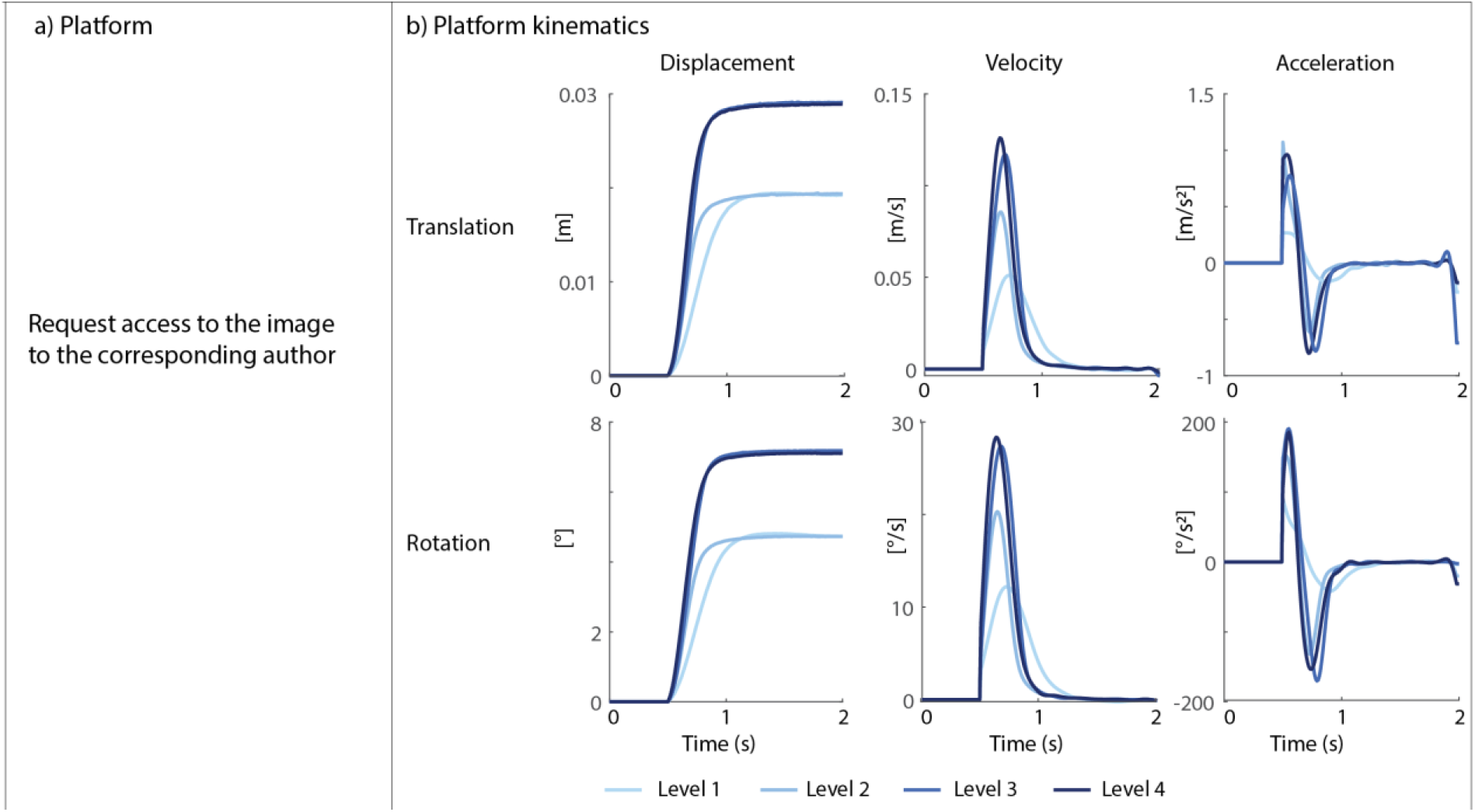
Reactive balance test set-up. **a)** Caren-platform for combined translational and rotational perturbations. Platform kinematics: (top) displacement, velocity, and acceleration of a point on the platform between the ankles and (bottom) angular displacement, velocity, and acceleration for the different perturbation levels (L1-L4).

### Protocol

For every participant, we collected age and anthropometrics (length and weight). We performed a clinical exam to determine the range of motion and Modified Ashworth Score for the children with CP. During balance assessment, participants stood barefoot on the platform. Before the start of the assessments, participants were instructed to stand upright and maintain balance without taking a step, unless necessary to avoid falling. When participants needed to take a step, we asked them to return their feet to the starting position, which was marked with tape on the platform. Arm movement was unconstrained. The protocol consisted of four increasingly difficult perturbation levels (increased platform displacement, velocity and/or acceleration, figure 1b). Within each perturbation level, eight perturbations were administered with 12s between perturbations. When the participant stepped in more than 3 out of 8 perturbations, we did not continue to the next level. If needed, rest was given between perturbation levels. The measurements described above were part of a larger single-session protocol more broadly assessing spasticity and balance.

### Data processing and analysis

Marker trajectories were processed using OpenSim 3.3 (18,19). A generic full-body musculoskeletal model (Gait2392 with arms, (20)) was scaled based on anatomic marker positions. Joint angles were computed using OpenSim’s Inverse Kinematics tool and CoM position was computed using OpenSim’s Body Kinematic tool. Anterior-posterior CoM displacement was expressed relative to the ankle and numerically differentiated to compute the CoM velocity. CoM acceleration was then computed using a Savitzky-Golay filter (21). Average ankle angles and CoM displacement, velocity, and acceleration were calculated across all non-stepping trials within each perturbation level for each participant and each level.

EMG data was band-pass filtered using a fourth order Butterworth filter between 10 and 450 Hz followed by signal rectification. Finally, a fourth order Butterworth low-pass filter with 40 Hz cut off was applied (22). The filtered EMG signal was scaled by the maximum value across all perturbations for every participant. As part of the bigger protocol, we also performed translational perturbations, which were also taken into account when determining the scaling value. The average EMG signal was calculated across all non-stepping trials for each participant and each level.

### Outcome parameters

#### a. Muscle activity, center of mass movement, and ankle kinematics

We computed average reactive activity LG, MG, SOL and TA in three time bins inspired by Golhover (23). The first time bin (Z1) started at platform onset and ended 150ms later, during this time interval we expected little reactive muscle activity due to the slow movement onset of the platform and neural transmission delay. The second time bin (Z2) lasted from 150ms to 250ms after platform onset, when most of the forward CoM movement occurred and balance correcting activity of the plantar flexors (LG, MG, SOL) was expected. The third time bin (Z3) lasted from 250ms to 400ms after platform onset when the CoM moved backward and balance correcting activity of the TA was expected (figure 2: row 5). Average reactive muscle activity was computed and subtracted by baseline activity (average muscle activity 100ms before perturbation onset) from the average filtered and scaled EMG signal (figure 2: bottom row).

**Figure 2:**
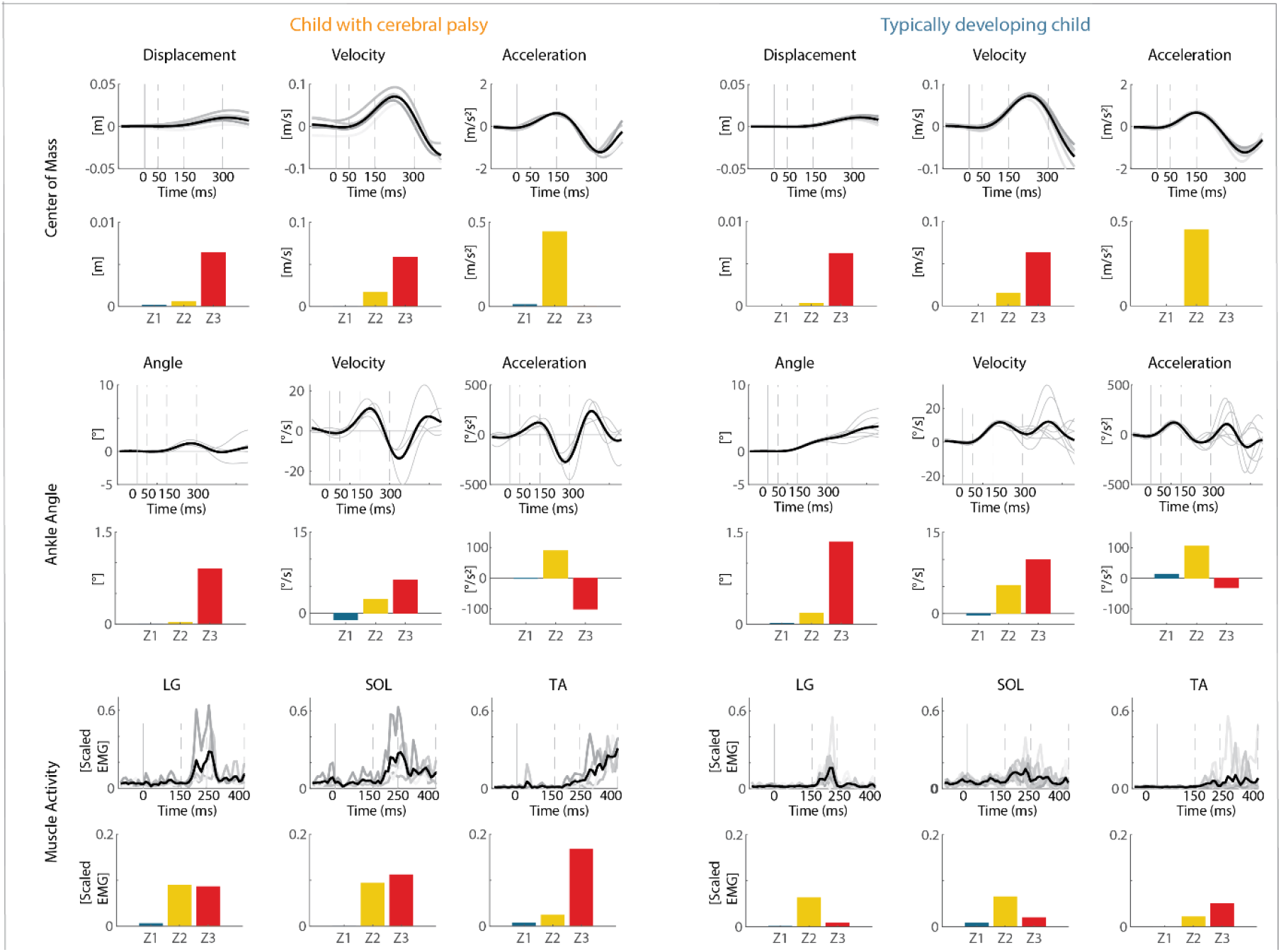
Exemplar cases for muscle activity, center of mass movement, and ankle kinematics for perturbation level 2 in time bins. Left panel: child with cerebral palsy, right panel: typically developing child. Row 1-2: Center of mass kinematics (displacement, velocity, and acceleration) as a function of time with indication of zones and average displacement, velocity, and acceleration per zone in blue for zone 1 (Z1), in yellow for zone 2 (Z2), and in red for zone 3 (Z3); Row 3-4: Ankle angle kinematics as a function of time with indication of zones and average ankle angular displacement, velocity, and acceleration per zone; Row 5-6: Muscle activations as a function of time with indications of zones and average muscle activity per zone. Light gray traces are separate trials within one level, black is the average across the level. Dotted lines indicate time zones. LG = lateral gastrocnemius; SOL = soleus; TA = tibialis anterior.

We similarly assessed the average CoM (figure 2: row 1-2) and ankle angle (figure 2: row 3-4) displacement, velocity, and acceleration. All time bins were shifted by 100ms, i.e., (i.e., Z1: onset platform to 50ms; Z2: 50ms to 150ms; Z3: 150ms to 300ms) as we assumed that the kinematic disturbances provided sensory inputs for the later muscle responses.

We assessed differences in average reactive muscle activity and kinematics for the three different zones between children with CP and TD children.

#### b. Co-contraction index

We calculated the co-contraction index (CCI) as the overlap in filtered and scaled EMG between TA and respectively LG, MG, and SOL (PF) (24):

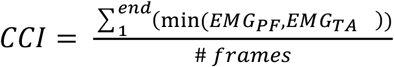

Low CCI values indicate that both muscles are active at different moments whereas high CCI values indicate that both muscles are active at the same time. We performed the analysis for two time intervals, i.e. between perturbation onset and 400ms after perturbation onset (interval covering the time bins described above), and between 0.5s before perturbation onset and 1.5s after perturbation onset (similar time interval as sensorimotor response model described below).

#### c. Sensorimotor response model

We tested the relation between muscle activity and CoM kinematics by reconstructing measured EMG trajectories by a weighted sum of delayed CoM acceleration, velocity, and displacement trajectories. We modeled both a balance correcting and an antagonistic feedback pathway (figure 3a). We used the following sensorimotor response model to reconstruct plantar flexor (LG, MG, SOL) activity (plantar flexors needed to restore upright position when CoM moved forward):

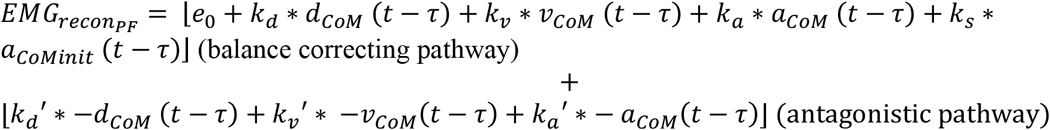

and the following sensorimotor response model to reconstruct tibialis anterior activity (tibialis anterior needed to restore upright position when CoM moved backward):

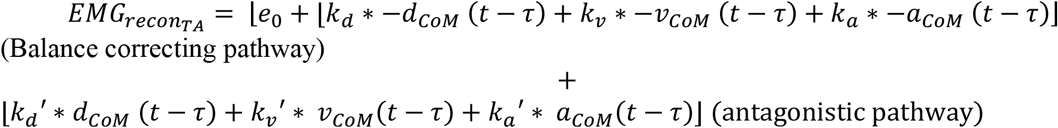

with *EMG*_*reconPF*_ and *EMG*_*reconTA*_ the reconstructed muscle activity for the plantar flexors and tibialis anterior, respectively; e_0_ baseline muscle activity; d_CoM_, v_CoM_, a_CoM_ center of mass displacement, velocity, and acceleration; k_d_, k_v_, k_a_ feedback gains or weights for the balance correcting pathway, k_d_’, k_v_’, k_d_’ feedback gains or weights for the antagonistic pathway, and a common time delay (τ) of 100 ms to account for processing and neural transmission time. We added a separate feedback term for the initial center of mass acceleration, a_CoM_init_, with a corresponding stiction gain, k_s_, inspired by Welch and Ting (9). The initial burst in EMG is proportional to the initial CoM acceleration and might be driven by the initial strong increase in spindle firing that coincides with short-range stiffness in the muscle (25,26). We therefore included this term until the change in the ankle angle was 0.5°, corresponding to the estimated short-range stiffness range (27). In contrast to Welch and Ting, we included both an acceleration and stiction term based on preliminary analyses. No stiction gain was used in the tibialis anterior muscle as in our protocol, this muscle was first shortened and short-range stiffness is strongly reduced with prior movement (26). Only the positive part of the signal (⌊.⌋) was used to represent excitatory drive to motor pools.

**Figure 3:**
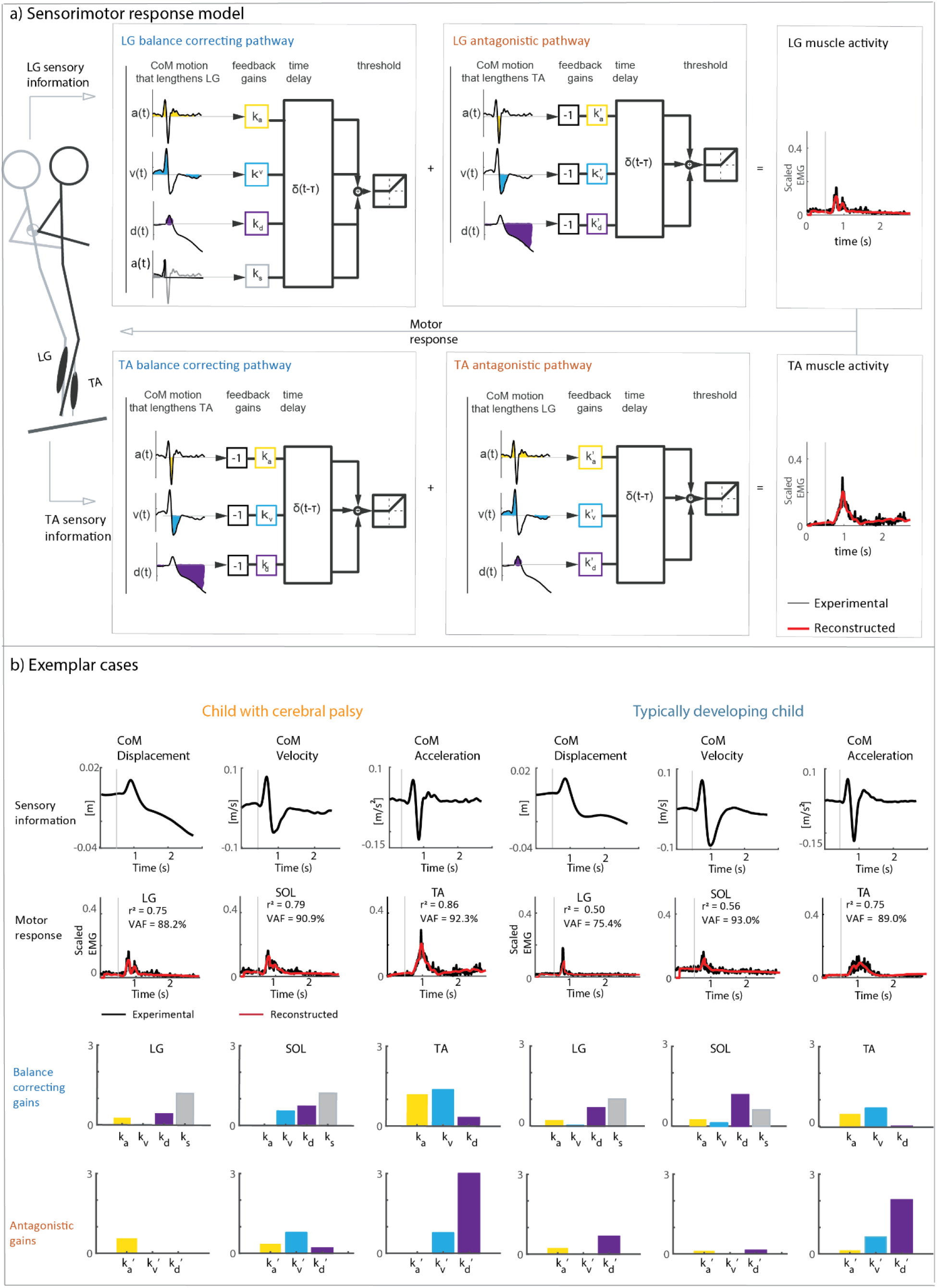
Sensorimotor response model. **a)** Graphical representation of the (extended) sensorimotor response model. Measured muscle activity (black) is reconstructed (red) using delayed feedback of CoM acceleration (yellow), velocity (blue), displacement (purple), and stiction (grey, for plantar flexors only) along a balance-correcting pathway (blue) and antagonistic pathway (orange; sensitive to CoM movement in the opposite direction). CoM acceleration, velocity, position and stiction are multiplied by subject specific feedback gains (balance correcting: k_a_, k_v_, k_d_, k_s_; antagonistic pathway: k_a_’, k_v_’, k_d_’). Note the opposite sign of the CoM kinematics in the balance correcting (blue) and antagonistic pathways (orange) of the plantar flexors (top row) and tibialis anterior (bottom row). In the extended model, both pathways are used (gains and prime gains), while in the simple model only the balance correcting pathway is used. **b)** Exemplar cases with the extended sensorimotor response model for one child with cerebral palsy (left) and one typically developing child (right). Top row: center of mass kinematics; second row: measured (black) and reconstructed (red) muscle activity signals; third row: balance correcting gains; bottom row: antagonistic gains (i.e., prime gains). LG = lateral gastrocnemius; SOL = soleus; TA = tibialis anterior.

Baseline activity (e_0_) was set to the measured data 0.5s before perturbation onset. Gains were estimated by minimizing the cost function, which was defined as the weighted sum of squared difference between reconstructed and measured EMG over a time interval from 0.5s before until 1.5s after perturbation onset (weight of 1) and squared prime gains (weight of 1E-4). Prime gains were penalized to discourage the use of the antagonistic pathway unless this had a considerable effect on the fit between reconstructed and measured EMG as we did not expect the antagonistic pathway to be active in TD children. All gains were constrained between 0 and 10/m (displacement, prime displacement), 10s/m (velocity and prime velocity), 10s^2^/m (acceleration, prime acceleration and stiction).

We assessed the goodness of fit between measured and reconstructed muscle activity (perturbation onset until 1.5s after perturbation onset) using the coefficient of determination (r^2^, calculated as the squared correlation coefficient), the variability accounted for (VAF, defined as the uncentered r^2^) and root mean square error (RMSE). Furthermore, we tested for differences in gains between children with CP and TD children (figure 3b). Different gains would indicate that the sensitivity to CoM perturbations differs between both groups of children.

We also explicitly tested whether EMG trajectories could be reconstructed without using the antagonistic feedback pathways (i.e., activity of the plantar flexors when the CoM moves backwards and activity of the tibialis anterior when the CoM moves forward) in both groups of children (figure 3a). We expected that the antagonistic pathway is not necessary to reconstruct reactive muscle activity in TD children, whereas adding the antagonistic pathway would improve the fit between reconstructed and measured EMG when reciprocal inhibition is impaired in children with CP. To test this, we calculated the difference in cost (squared difference between reconstructed and measured activations) between both models (with prime gains (extended model) and without prime gains (simple model)). We expressed this improvement as a percentage of the cost of the simple model.

### Statistical analysis

All statistical analyses were performed using Matlab (R2018b, Mathworks, United States) with differences considered significant at p < 0.05.

Differences in average muscle activity (for LG, MG, SOL, TA), center of mass kinematics (displacement, velocity, and acceleration) and ankle angle kinematics (change, angular velocity, or angular acceleration) between children with CP and TD children were tested for each perturbation level using a linear mixed model with two fixed effects: (1) group (CP vs. TD) and (2) time bin (1 to 3, ordinal). A participant factor was included as a random factor nested within group.

The co-contraction index was compared between children with CP and TD children using a linear mixed model with two fixed effects: (1) group (CP vs. TD) and (2) perturbations level (1 to 4, ordinal). A participant factor was included as a random factor nested within group.

Differences in gains between groups were tested using a linear mixed model with two fixed effects: (1) group (CP vs. TD) and (2) perturbation level (1 to 4, ordinal) for each muscle separately. A participant factor was included as a random factor nested within group. All tests for differences in gains for each muscle were performed without adjustments of simultaneous interference as they were assumed to evaluate independent hypothesis.

Improvements in cost for the extended model compared to the simple model were compared between children with CP and TD children using a linear mixed model with two fixed effects: (1) group (CP vs. TD) and (2) perturbation level (1 to 4, ordinal). A participant factor was included as a random factor nested within group.

## RESULTS

Due to technical errors in EMG recordings, we had to exclude data of LG in one TD child, SOL in one child with CP, and TA in another child with CP.

All TD children performed all levels. Four children with CP did not perform levels 2-4 and one child with CP did not perform levels 3-4.

### a) Muscle activity, center of mass movement, and ankle kinematics

#### Muscle activity

Children with CP modulated their muscle response differently in time than TD children, indicating a reduced ability to switch between plantar flexors and tibialis anterior upon reversal of the CoM movement (figure 4, supplementary material S3 (table S2)). Muscle activity was different between time bins and there was an interaction effect between group and time bin for most muscles and perturbation levels (LG: L1, L2, L3, p < 0.04; MG: L2, L3, p < 0.02; SOL: L1, L2, L3, p < 0.02, TA: L1, L2,

**Figure 4:**
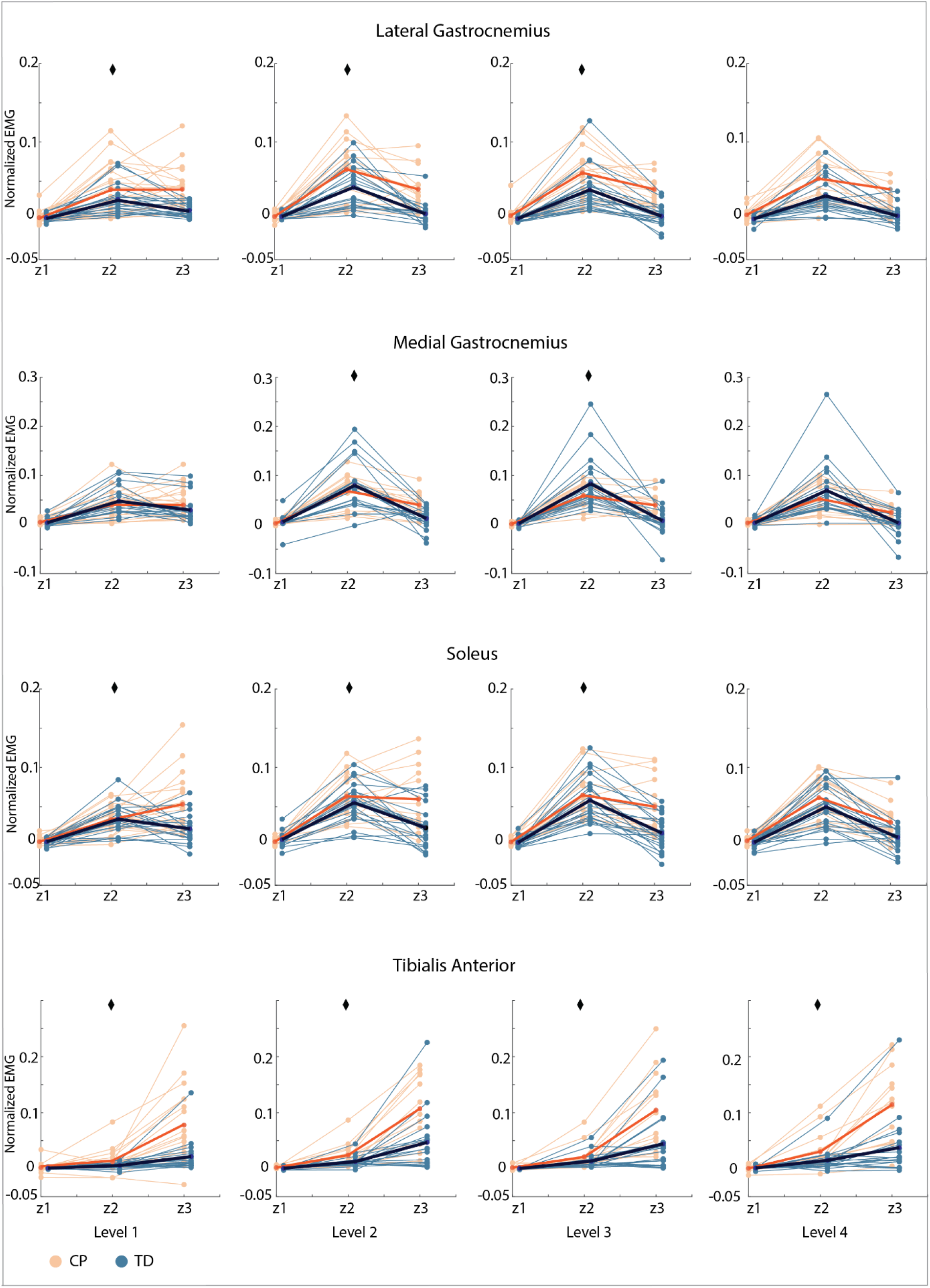
Average normalized EMG for three time bins (Z1-Z3) for all muscles and all levels. Children with cerebral palsy (CP) in orange, typically developing (TD) children in blue. Group average for children with cerebral palsy in red, group average for typically developing children in dark blue. Significant interaction effects between time bin and group (p < 0.05) are indicated with a diamond.

L3, L4, p < 0.005). Visual inspection revealed that the reduction in plantar flexor activity from the second (little average CoM displacement) to the third time bin (average forward CoM displacement) that was present in TD children was reduced or even absent in children with CP. Furthermore, the increase in tibialis anterior muscle activity from the second to the third time bin was higher in children with CP than in TD children.

#### Center of mass movement

CoM displacement, velocity, and acceleration was different between time bins but not between groups, indicating that there were no differences in CoM movement between groups (figure 5a, supplementary material S3 (table S3)).

**Figure 5:**
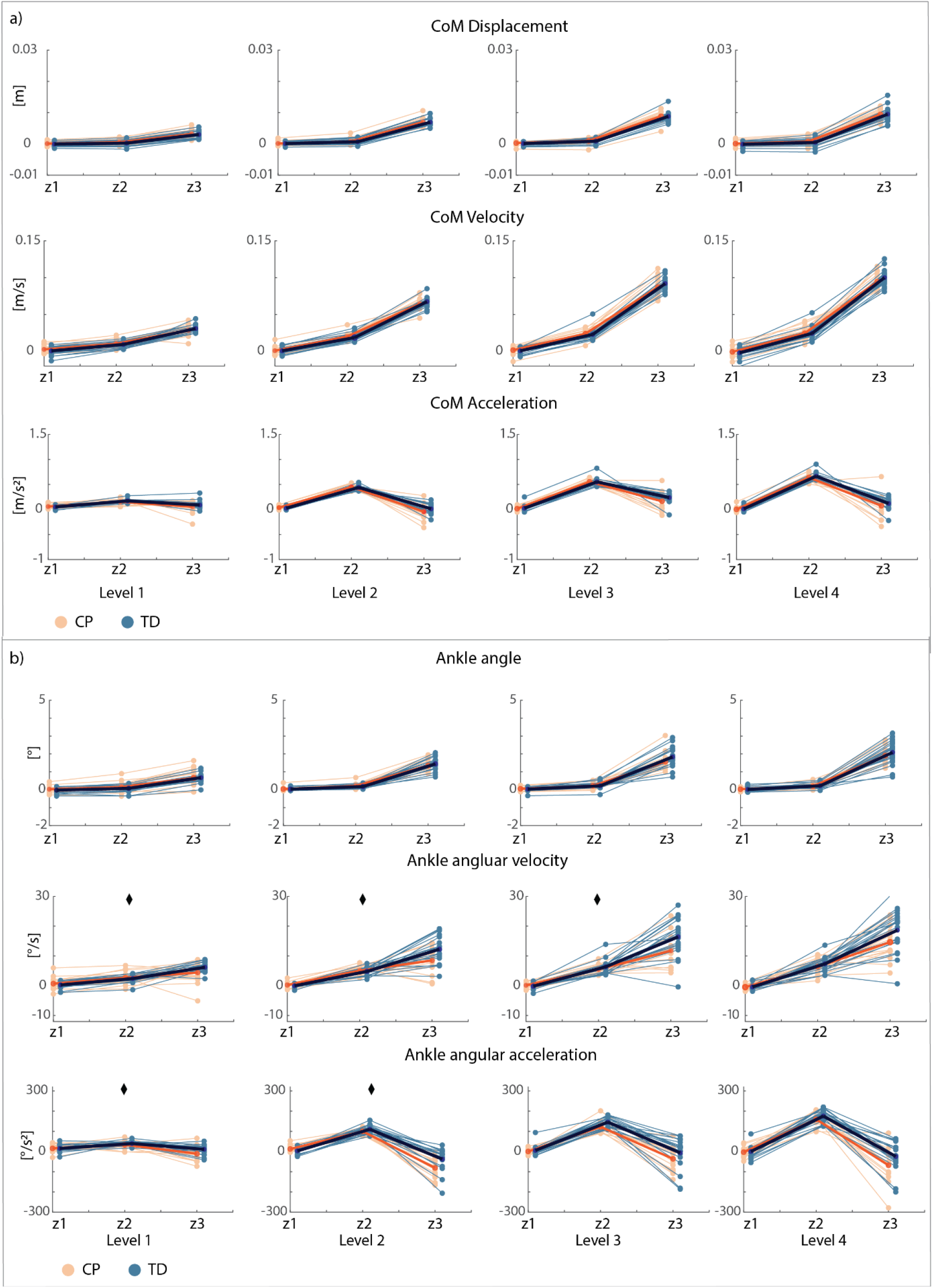
Average center of mass (**a**) and ankle kinematics (**b**) for three time bins (Z1-Z3) for all levels. Children with cerebral palsy (CP) in orange, typically developing (TD) children in blue. Group average for children with cerebral palsy in red, group average for typically developing children in dark blue. Significant interaction effects between time bin and group (p < 0.05) are indicated with a diamond.

#### Ankle kinematics

Ankle displacement was different between time bins but not between groups. Ankle angular velocity and acceleration were different between time bins and there was an interaction effect between time bin and group for most perturbation levels (velocity: L1, L2, L3, p < 0.02; acceleration: L1, L2, p < 0.05). Visual inspection reveals a lower angular velocity and larger angular deceleration in the last time bin in children with CP (figure 5b, supplementary material S3 (table S4)).

### b) Co-contraction index

Co-activation was higher in children with CP than in TD children over the entire response. The CCI was significantly higher (p<0.001) for children with CP than for TD children across all perturbation levels and for all muscle pairs (LG and TA, MG and TA, SOL and TA) for both the shorter (0-400ms after perturbation onset) (figure 6, supplementary material S4 (table S5)) and longer (0.5s before until 1.5s after perturbation onset) time interval (supplementary material S4 (figure S2, table S6)).

**Figure 6:**
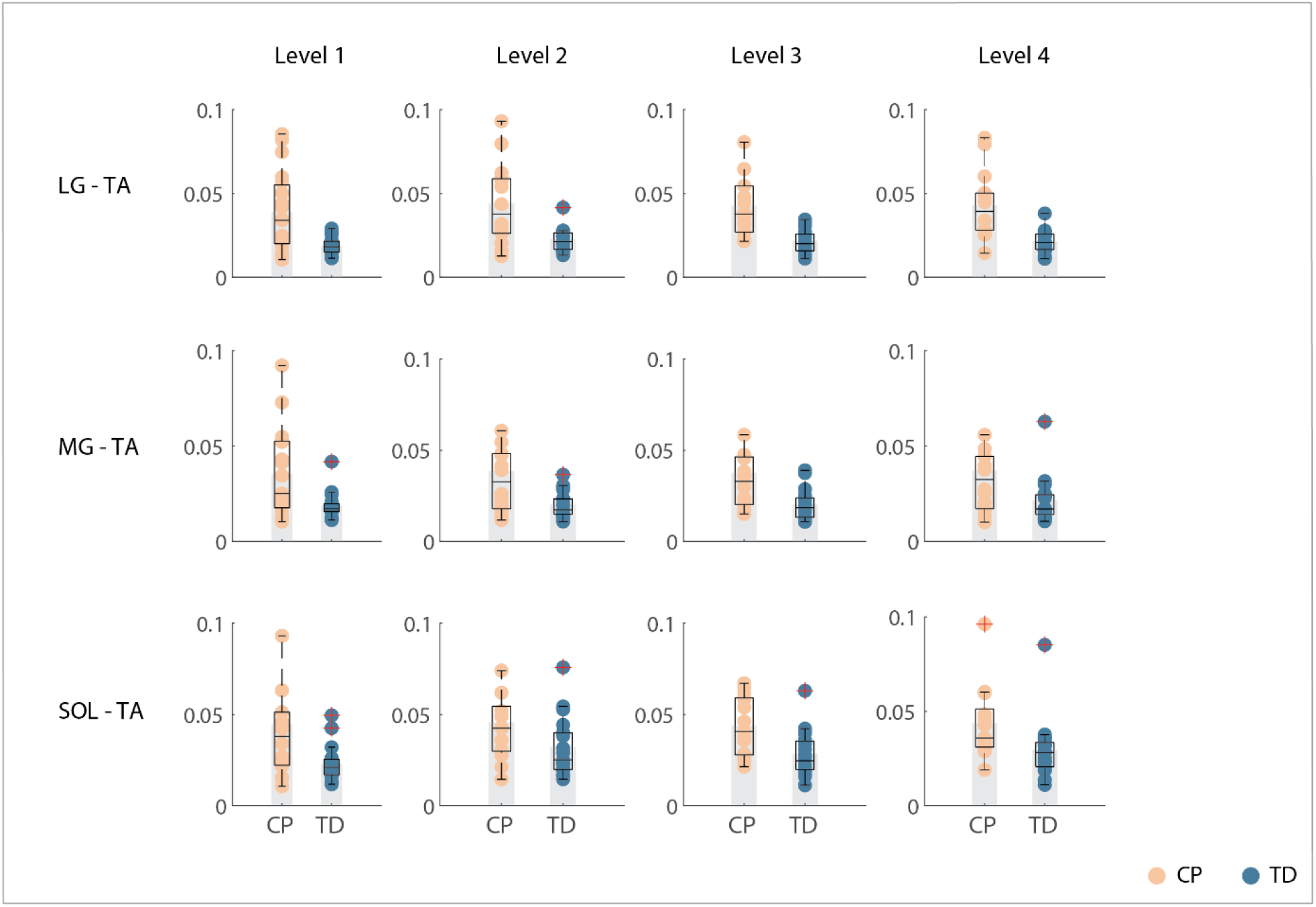
Co-contraction index for perturbation onset until 350ms after perturbation onset. Children with cerebral palsy (CP) in orange, typically developing (TD) children in blue. Grey bars indicate group averages, boxplots in black indicate median and interquartile range and dots represent individual scores. Groups are significantly different across all levels for all muscle pairs. LG = lateral gastrocnemius; MG = medial gastrocnemius; SOL = soleus; TA = tibialis anterior.

### c) Sensorimotor response model

Muscle EMG during combined translational and rotational perturbations of standing can be reconstructed by delayed CoM feedback in both children with CP and TD children as reflected in the high goodness of fit values (r^2^ - CP: 0.60 ± 0.21, TD 0.50 ± 0.20; VAF – CP: 90.04% ± 6.9%, TD: 86.9% ± 7.1%) and low error scores (RMSE – CP: 0.018 ± 0.01, TD: 0.015 ± 0.01) (figure 7a, supplementary material S5 (table S7)).

**Figure 7:**
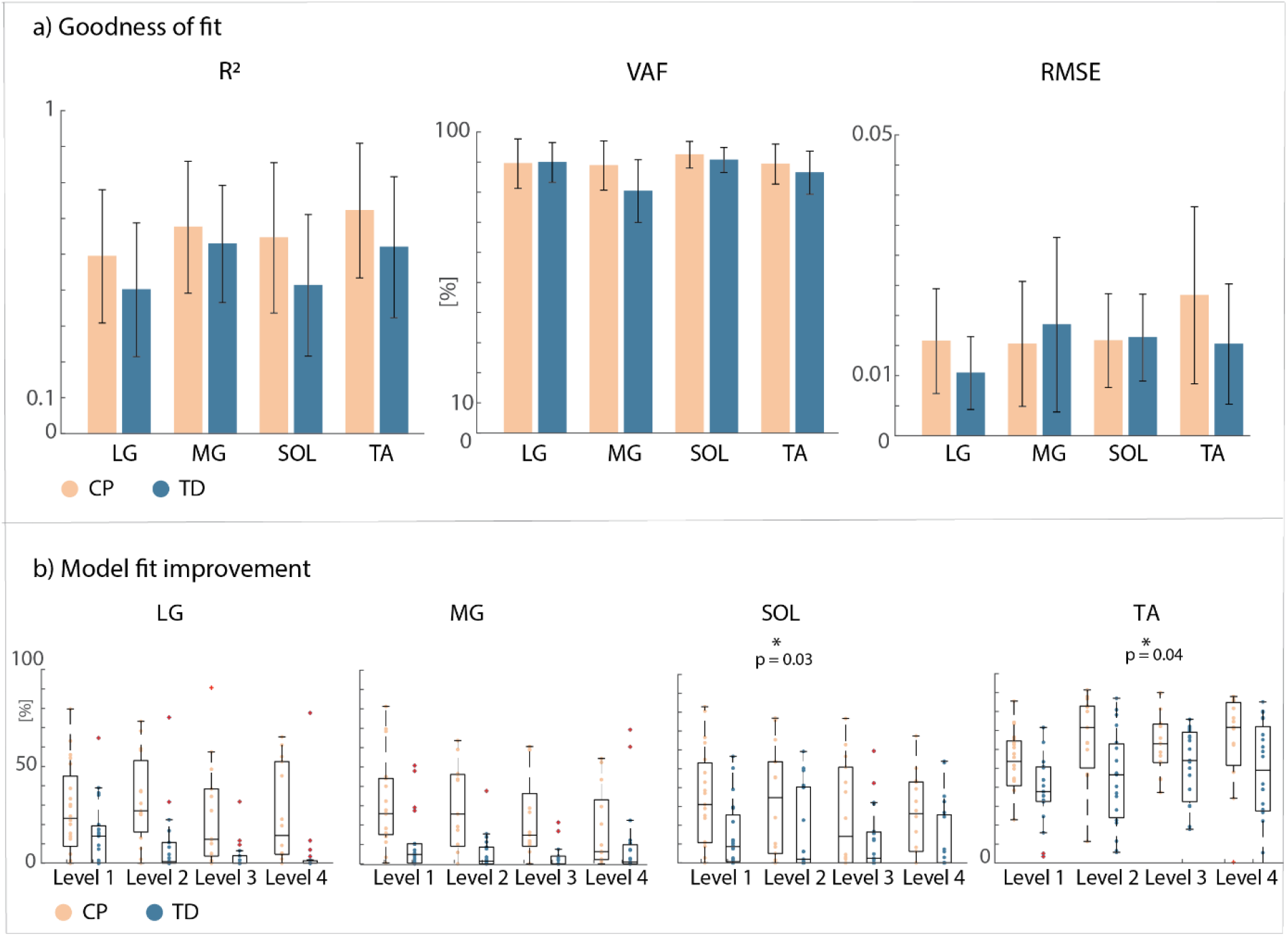
Goodness of fit for the extended sensorimotor response model and reduction in cost for the extended versus simple sensorimotor response model. **a)** Goodness of fit values and error scores across all levels for all muscles for the extended (with antagonistic pathways) sensorimotor response model. Error bars are indicated in black. r^2^ = r squared; VAF = variance accounted for; RMSE = root mean square error. **B)** Improvement in cost (%) when using the extended versus simple (no antagonistic pathways) sensorimotor response model for all muscles. The cost is the squared difference between measured and reconstructed EMG. Boxplots in black indicate median and interquartile range, dots represent individual scores. Significant differences between groups (CP vs. TD) are indicated with a star and p-values. LG = lateral gastrocnemius; MG = medial gastrocnemius; SOL = soleus; TA = tibialis anterior. Children with cerebral palsy (CP) in orange, typically developing (TD) children in blue.

The fitting error (defined as the squared error between measured and reconstructed signals) was lower when adding the antagonistic pathways for both children with CP (LG: 26.4% ± 3.7%; MG: 23.4% ± 4.5%; SOL: 26.0% ± 8.5%; TA: 63.2% ± 6.8%) and TD children (LG: 8.2% ± 4.5%; MG: 5.5% ± 2.6%; SOL: 12.1% ± 3.8%; TA:43.9% ± 5.1%) (figure 7b, supplementary material S5 (table S8)). Adding the antagonistic pathway induced a larger reduction in the fitting error for children with CP than for TD children for the soleus (p = 0.034) and tibialis anterior (p = 0.044) across all levels. Children with CP had higher gains for CoM feedback than TD children for all muscles (figure 8, supplementary material S5 (figure S3, table S9)).

**Figure 8:**
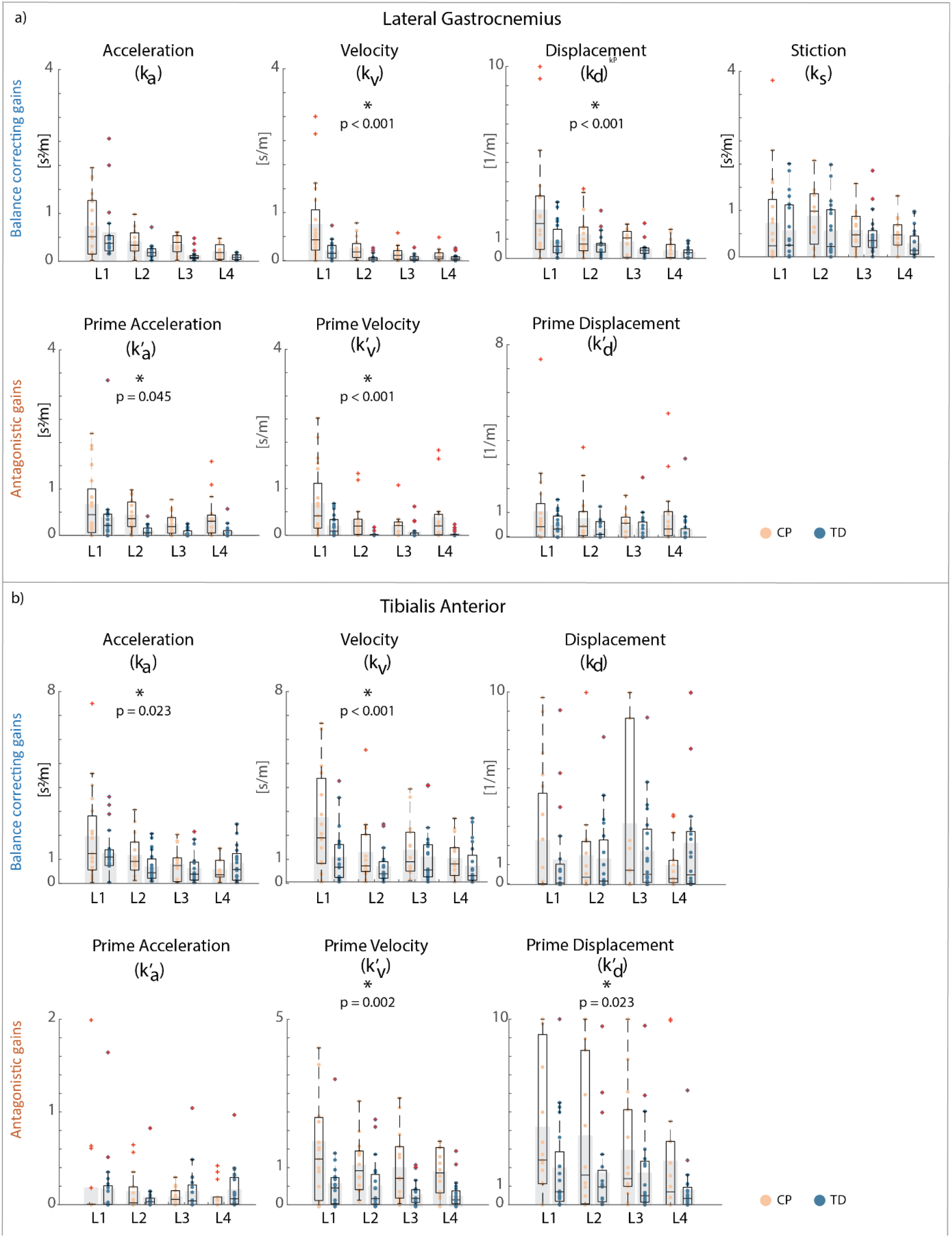
Center of mass feedback gains between children with cerebral palsy and typically developing children. a) Lateral gastrocnemius. **B)** Tibialis Anterior. Differences between children with CP and TD children were larger for lateral gastrocnemius than for soleus and medial gastrocnemius, therefore results for lateral gastrocnemius are shown in the main figure. Figures for medial gastrocnemius and soleus can be found in supplementary material S5 (figure S3). Upper row: balance correcting pathway gains, bottom row: antagonistic pathway gains. L1-L4: Level 1 to level 4. Grey bars indicate group average, boxplots in black indicate mean and interquartile ranges, and dots represent individual scores. Children with cerebral palsy (CP) in orange, typically developing (TD) children in blue. Significant differences (p<0.05) between groups are indicated with a star and p-values.

For the lateral gastrocnemius, velocity (222%, p < 0.0001), displacement (109%, p = 0.0001), prime acceleration (186%, p = 0.045), and prime velocity (382%, p < 0.0001) gains were higher in children with CP than in TD children across all levels (figure 8a). Furthermore, there was an interaction effect between level and group for velocity (p = 0.005) and displacement (p = 0.039) gains suggesting a larger difference between both groups for the lower levels (visual exploration). All gains, except for the prime displacement gain decreased with increasing level (p < 0.005).

For the medial gastrocnemius, velocity (74%, p = 0.002), prime acceleration (213%, p < 0.001), and prime velocity (973%, p < 0.001) gains were higher in children with CP than in TD children across all levels (figure S3a). The acceleration gain was lower for children with CP than for TD children (7%, p = 0.004). Furthermore, there was an interaction effect between level and group for acceleration (p = 0.005) and stiction (p = 0.025) gains. Acceleration, velocity, displacement, prime acceleration, and prime velocity gains decreased with increasing perturbation level (p < 0.05).

For the soleus, acceleration (42%, p = 0.018), velocity (124%, p = 0.016), displacement (32%, p = 0.020), prime acceleration (142%, p < 0.001), and prime velocity (136%, p = 0.003) gains were higher in children with CP than in TD children across all levels (figure S3b). No interaction effect between group and level was observed. All gains, except for prime displacement and stiction gains, decreased with increasing levels (p < 0.05).

For the tibialis anterior, acceleration (35%, p = 0.023), velocity (85%, p < 0.001), prime velocity (175%, p = 0.002), and prime displacement (113%, p = 0.022) gains were higher in children with CP than in TD children across all levels (figure 8b). Furthermore, there was an interaction effect between level and group for the velocity gain (p = 0.012), suggesting larger differences between groups for the lower levels (visual exploration). Acceleration and velocity gains decreased with increasing level (p < 0.001).

## DISCUSSION

Reduced reciprocal inhibition might contribute to balance impairments in CP. We perturbed standing balance by a combined translational and rotational movement of the support surface that induced a similar initial forward CoM movement followed by a backward CoM movement in children with CP and TD children. In TD children, we observed activation of the plantar flexors followed by activation of the tibialis anterior. This switch from plantar flexor to tibialis anterior activity was less pronounced in children with CP (figure 4). The reduced ability to selectively activate muscles in response to CoM movement was reflected in increased co-activation of plantar flexors and tibialis anterior throughout the response and in the gains of the sensorimotor response model. In both children with CP and TD children, muscle activity in response to combined translational and rotational perturbations could be explained by delayed CoM feedback. However, we found that accounting for antagonistic pathways was more needed in children with CP than in TD children to reconstruct measured EMG. In addition, feedback gains of both the balance-correcting and antagonistic pathways were higher in children with CP than in TD children. Higher joint stiffness, as induced by muscle co-activation, hinders balance control in response to rotational perturbations where retaining an upright posture depends on the ability to dissociate the motion of the body from the motion of the feet. Our results thus suggest that muscle co-activation in CP is due to neural impairments rather than being a compensation strategy during balance perturbations.

Decreased reciprocal inhibition is a common symptom in children with CP, contributing to excessive antagonistic muscle co-activation during voluntary movement such as walking (5,6,28). Although increased muscle co-activation was observed in response to translational perturbations of standing balance in CP, this was not tested before during rotational perturbations. Our results show that upon reversal of the CoM movement, the tibialis anterior muscle becomes active – as expected -, but the expected reciprocal inhibition of the plantar flexors is lacking in children with CP, while a decrease in plantar flexor activity is clearly observed in TD children.

Our observation that delayed CoM feedback could explain muscle activity in response to combined rotational and translational perturbations suggests that task-level feedback is preserved in children with CP. We previously demonstrated that CoM feedback could explain muscle activity in response to support surface translations in children with CP. Since support surface translations elicit correlated CoM and ankle movements, we could not confidently conclude that task-level feedback was preserved in CP. Combined backward translational and toe-up rotational perturbations induce a change in the direction of the CoM displacement (forward in response to translation followed by backward in response to rotation) while the ankle continues to dorsiflex. Using such combined translational and rotational perturbations, it was demonstrated that muscle activity is dictated by CoM movement in healthy adults (15,16,23). Healthy adults first activate their gastrocnemius and then switch to activating their tibialis anterior (16,23). Although these experiments demonstrated that reactive muscle activity is driven by CoM movement rather than by joint movement during combined translational and rotational perturbations, we tested this for the first time – to our knowledge –using an explicit sensorimotor response model in both TD children and children with CP.

We are unable to attribute alterations in responsive muscle activity to local or task-level feedback pathways. Both spinal and supraspinal feedback pathways are involved in reactive balance (29) and both pathways are impaired in CP. Children with CP often have reflex hyper-excitability reflecting spinal feedback deficits (7,30). In addition, they have difficulties with selectively controlling their muscles due to impaired supraspinal pathways (7,30). Reflex hyper-excitability might lead to exaggerated responses of both the plantar flexors, which are stretched during balance perturbations, and the antagonistic tibialis anterior. Such increased sensitivity to muscle stretch might prevent children with CP to suppress plantar flexor activity when the CoM starts to move backwards, requiring compensations in supraspinal control to stabilize posture. Alternatively, the reduced ability to selectively activate muscles might rely in the supraspinal pathways. To gain more insight in the underlying pathways, we explored the relation between the gains and a clinical score of joint hyper-resistance, i.e., the MAS score of the gastrocnemius (exploratory analysis, supplementary material S6 (figure S4-S5)). We did not find any correlations. However, this might be due to inherent limitations of the MAS score (31). For example, the MAS score does not distinguish contributions from reflex hyper-excitability and altered passive tissue properties to joint hyper-resistance. A more in-depth analysis is thus required.

Notwithstanding the altered control of reactive balance, CoM displacements were not larger in children with CP than in TD children. We found that the maximum CoM excursion in response to translational perturbations was smaller in children with CP than in TD children (4). This might indicate that co-activation indeed helps to maintain an upright position when standing is perturbed by translational perturbations. Here, we found no differences in maximum CoM excursion between both groups. This suggest that children with CP were able to compensate for the increased co-activation when controlling the CoM position in response to rotational perturbations. We observed differences in the ankle angle velocity and acceleration, suggesting less ankle dorsiflexion movement. Children with CP thus likely compensated in other joints to control their CoM position.

### Conclusion

Reduced reciprocal inhibition might underlie altered standing balance control in children with CP, leading to increased co-activation in response to both support surface translations and rotations. Notwithstanding this reduced reciprocal inhibition, task-level control of the CoM is preserved in the rather functional (GMFCS I and II) children with CP included in this study. Future work should explore whether alterations in balance control are also correlated with falls.

## Data Availability

All data produced in the present study are available upon reasonable request to the authors

