## Supplementary material for "Combined translational and rotational perturbations of standing balance reveal contributions of reduced reciprocal inhibition to balance impairments in children with cerebral palsy"

### SUPPLEMENT

#### S1. Additional information on children with cerebral palsy.

Table S1: Additional information on children with cerebral palsy

| | GMFCS | Hemi vs. Di | Tested leg | MAS (LG/MG) |
|---|---|---|---|---|
| **CP1** | 1 | D | R | 1 |
| **CP2** | 1 | H | L | 1+ |
| **CP3** | 1 | H | L | 1 |
| **CP4** | 2 | H | R | 3 |
| **CP5** | 2 | H | R | 0 |
| **CP6** | 2 | D | R | 1+ |
| **CP7** | 1 | H | L | 3 |
| **CP8** | 1 | H | R | 1 |
| **CP9** | 1 | H | R | 1 |
| **CP10** | 1 | D | R | 0 |
| **CP11** | 1 | H | L | 1+ |
| **CP12** | 1 | H | L | 1 |
| **CP13** | 1 | H | L | 0 |
| **CP14** | 2 | D | L | 1 |
| **CP15** | 1 | H | R | 1 |
| **CP16** | 1 | H | R | 1 |
| **CP17** | 1 | H | R | 1 |
| **CP18** | 1 | D | L | 1 |
| **CP19** | 2 | H | R | 1+ |
| **CP20** | 1 | D | L | 0 |

GMFCS = Gross Motor Function Classification System (range 1-5)
H = Hemiplegic; D = diplegic; L = left; R = right;
MAS = Modified Ashworth Scale for the gastrocnemii (range 1-4)

**S2. Full body marker set-up.**

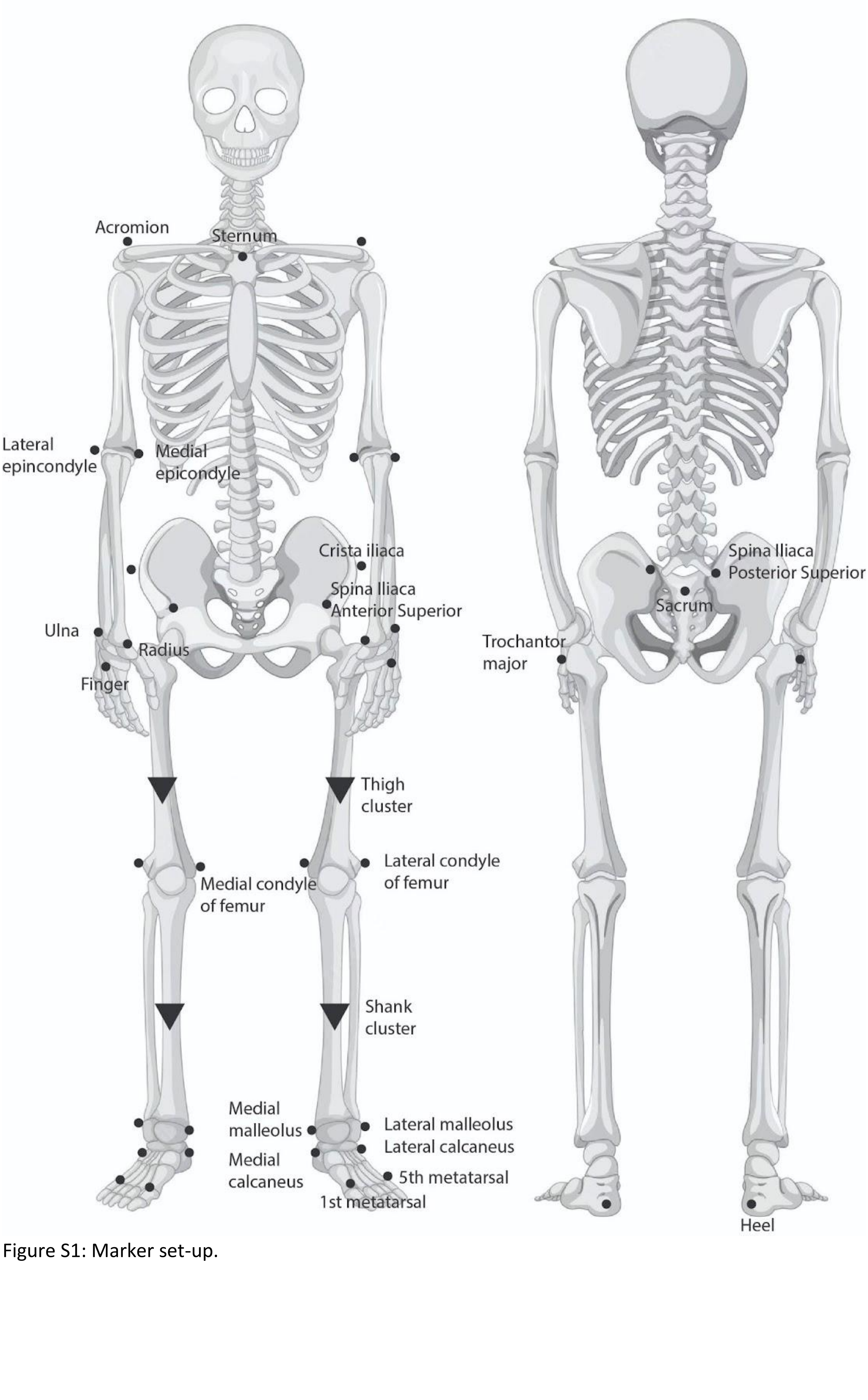


Figure S1: Marker set-up.

**S3. Muscle activity, center of mass movement, and ankle kinematics**

3.1. Muscle activity

Table S2: Statistical outcome parameters (p-values) for EMG zones for children with cerebral palsy and typically developing children.

| LG | p-value | | |
|---|---|---|---|
| | Time bin | Group | Bin: Group |
| Level 1 | **p < 0.001** | 1.00 | **0.013** |
| Level 2 | **p < 0.001** | 1.00 | **0.020** |
| Level 3 | **p < 0.001** | 0.59 | **0.034** |
| Level 4 | **p < 0.001** | 0.39 | 0.127 |

| TA | p-value | | |
|---|---|---|---|
| | Tim bin | Group | Bin: Group |
| Level 1 | **p < 0.001** | 0.91 | **p < 0.001** |
| Level 2 | **p < 0.001** | 0.90 | **0.001** |
| Level 3 | **p < 0.001** | 0.90 | **0.002** |
| Level 4 | **p < 0.001** | 0.96 | **p < 0.001** |

| MG | p-value | | |
|---|---|---|---|
| | Time bin | Group | Bin: Group |
| Level 1 | **p < 0.001** | 0.91 | 0.075 |
| Level 2 | **p < 0.001** | 0.79 | **0.022** |
| Level 3 | **p < 0.001** | 0.92 | **0.002** |
| Level 4 | **p < 0.001** | 0.97 | 0.051 |

| SOL | p-value | | |
|---|---|---|---|
| | Time bin | Group | Bin: Group |
| Level 1 | **p < 0.001** | 0.99 | **0.001** |
| Level 2 | **p < 0.001** | 0.75 | **0.006** |
| Level 3 | **p < 0.001** | 0.94 | **0.013** |
| Level 4 | **p < 0.001** | 0.79 | 0.254 |

LG = lateral gastrocnemius; MG = medial gastrocnemius; SOL = soleus; TA = tibialis anterior.
Significant differences (p<0.05) are indicated in bold.

3.2. Center of mass movement

Table S3: Statistical outcome parameters (p-values) for CoM zones for children with cerebral palsy and typically developing children.

| **Displacement** | p-value | | |
|---|---|---|---|
| | Time bin | Group | Bin: Group |
| Level 1 | **p < 0.001** | 0.60 | 0.94 |
| Level 2 | **p < 0.001** | 0.95 | 0.99 |
| Level 3 | **p < 0.001** | 0.69 | 0.97 |
| Level 4 | **p < 0.001** | 0.68 | 0.86 |
| **Velocity** | **p-value** | | |
| | Time bin | Group | Bin: Group |
| Level 1 | **p < 0.001** | 0.21 | 0.14 |
| Level 2 | **p < 0.001** | 0.93 | 0.29 |
| Level 3 | **p < 0.001** | 0.71 | 0.18 |
| Level 4 | **p < 0.001** | 0.69 | 0.24 |
| **Acceleration** | **p-value** | | |
| | Time bin | Group | Bin: Group |
| Level 1 | **p < 0.001** | 0.68 | 0.30 |
| Level 2 | **p < 0.001** | 0.67 | 0.13 |
| Level 3 | **p < 0.001** | 0.75 | 0.19 |
| Level 4 | **p < 0.001** | 0.78 | 0.79 |

Significant differences are indicated in bold.

3.3. Ankle kinematics

Table S4: Statistical outcome parameters (p-values) for ankle angle kinematics for children with cerebral palsy and typically developing children.

| **Position** | **p-value** | | |
|---|---|---|---|
| | Time bin | Group | Bin: Group |
| Level 1 | **$p < 0.001$** | 0.53 | 0.73 |
| Level 2 | **$p < 0.001$** | 0.95 | 0.40 |
| Level 3 | **$p < 0.001$** | 0.85 | 0.12 |
| Level 4 | **$p < 0.001$** | 0.91 | 0.46 |
| **Velocity** | **p-value** | | |
| | Time bin | Group | Bin: Group |
| Level 1 | **$p < 0.001$** | 0.46 | **0.01** |
| Level 2 | **$p < 0.001$** | 0.77 | **$p < 0.001$** |
| Level 3 | **$p < 0.001$** | 0.81 | **0.01** |
| Level 4 | **$p < 0.001$** | 0.96 | 0.10 |
| **Acceleration** | **p-value** | | |
| | Time bin | Group | Bin: Group |
| Level 1 | **$p < 0.001$** | 0.98 | **0.05** |
| Level 2 | **$p < 0.001$** | 0.42 | **0.01** |
| Level 3 | **$p < 0.001$** | 0.73 | 0.49 |
| Level 4 | **$p < 0.001$** | 0.85 | 0.21 |

Significant differences are indicated in bold.

**S4. Co-contraction index**

Table S5: Statistical outcome parameters (p-values) for co-contraction index between children with cerebral palsy and typically developing children for the time frames similar as the time bins (onset-350ms after onset).

| CCI - short | p-values | | |
|---|---|---|---|
| | Level | Group | Group:Level |
| LG – TA | **0.0005** | **p < 0.001** | 0.34 |
| MG – TA | **0.0017** | **0.005** | 0.15 |
| SOL - TA | 0.0984 | **0.0020** | 0.99 |

LG = lateral gastrocnemius; MG = medial gastrocnemius; SOL = soleus; TA = tibialis anterior.
Significant differences are indicated in bold.

Table S6: Statistical outcome parameters (p-values) for co-contraction index between children with cerebral palsy and typically developing children for the time frames similar as the sensorimotor response model.

| CCI - long | p-values | | |
|---|---|---|---|
| | Level | Group | Level:Group |
| LG - TA | 0.14 | **p < 0.001** | 0.66 |
| MG - TA | **0.02** | **p < 0.001** | 0.11 |
| SOL - TA | 0.10 | **0.001** | 0.53 |

LG = lateral gastrocnemius; MG = medial gastrocnemius; SOL = soleus; TA = tibialis anterior.
Significant differences are indicated in bold.

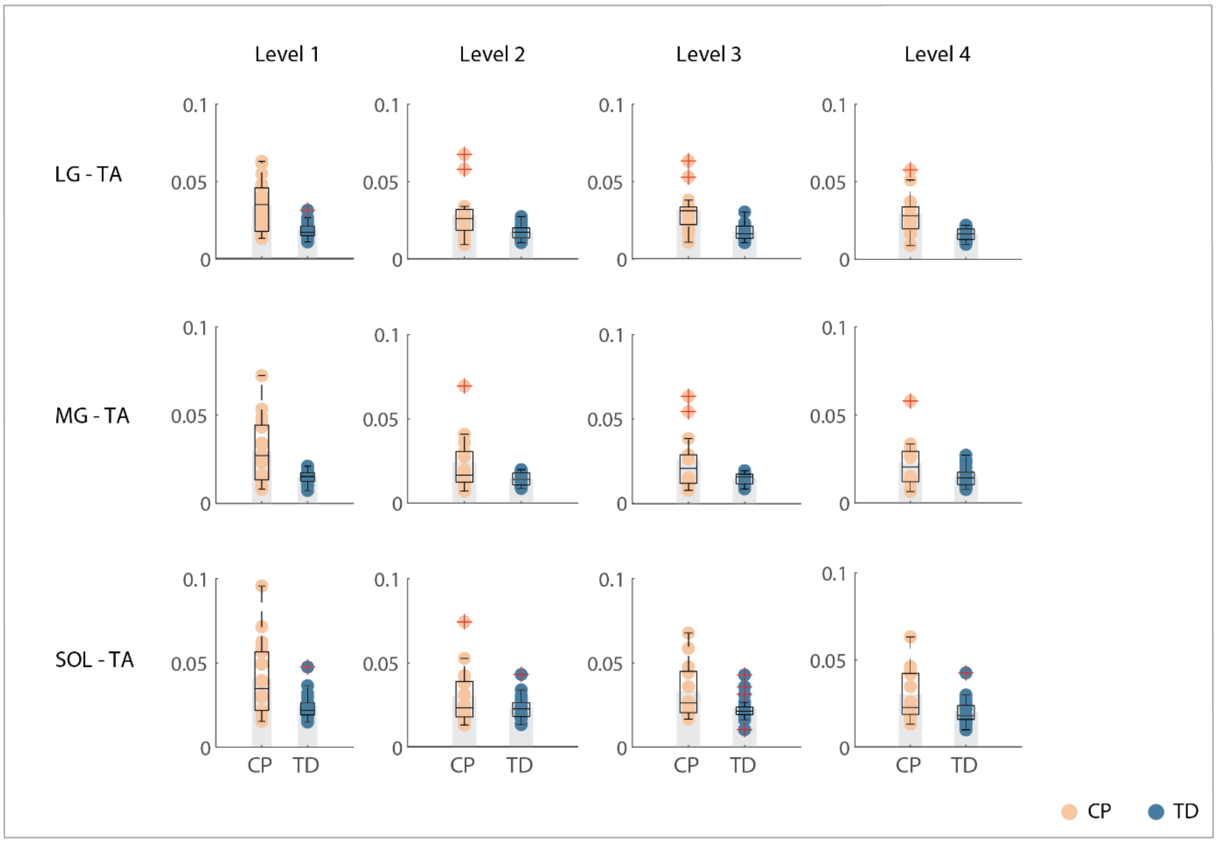


Figure S2: Co-contraction index for 0.5s before perturbation onset until 1.5s after perturbation onset. Children with cerebral palsy (CP) in orange, typically developing (TD) children in blue. Grey bars indicate group averages, boxplots in black indicate median and interquartile range and dots represent individual scores. Groups are significantly different across all levels for all muscle pairs.

LG = lateral gastrocnemius; MG = medial gastrocnemius; SOL = soleus; TA = tibialis anterior.

**S5. Sensorimotor response model**

5.1. Goodness of fit values

Goodness of fit values are high for both children with cerebral palsy and typically developing children.

Table S7: Goodness of fit and error scores (mean and standard deviations) for the extended sensorimotor response model for children with cerebral palsy and typically developing children.

| **Goodness of fit** | **CP** mean | (± SD) | **TD** mean | (± SD) |
|---|---|---|---|---|
| | $R^2$ | | | |
| LG | 0.53 | 0.20 | 0.43 | 0.20 |
| MG | 0.62 | 0.20 | 0.57 | 0.18 |
| SOL | 0.59 | 0.22 | 0.44 | 0.21 |
| TA | 0.67 | 0.20 | 0.56 | 0.21 |
| | VAF [%] | | | |
| LG | 89.52 | 8.23 | 89.91 | 6.61 |
| MG | 88.86 | 8.21 | 80.39 | 10.39 |
| SOL | 92.44 | 4.44 | 90.70 | 4.14 |
| TA | 89.33 | 6.63 | 86.47 | 7.14 |
| | RMSE | | | |
| LG | 0.016 | 0.009 | 0.010 | 0.006 |
| MG | 0.015 | 0.010 | 0.019 | 0.015 |
| SOL | 0.016 | 0.008 | 0.016 | 0.007 |
| TA | 0.023 | 0.015 | 0.015 | 0.010 |

$R^2$: R-squared, indicating fit with overall pattern; VAF= variance accounted for, indicating fit with amplitude of response activity; RMSE = root mean square error, indicating absolute error between measured and reconstructed signal
LG = lateral gastrocnemius; MG = medial gastrocnemius; SOL = soleus; TA = tibialis anterior
CP = cerebral palsy; TD = typically developing

5.2. Improvement in fit when adding antagonistic muscle pathways

Table S8: Improvement in fit (mean and standard deviations) when adding antagonistic feedback pathways (extended model vs. simple model) for children with cerebral palsy and typically developing children.

| **Fit improvement [%]** | **CP** Mean | (± SD) | **TD** Mean | (± SD) | **p-value** Level | Group | Level:Group |
|---|---|---|---|---|---|---|---|
| **LG** | 26.4 | 3.7 | 8.2 | 4.5 | 0.33 | 0.24 | 0.15 |
| **MG** | 23.4 | 4.5 | 5.5 | 2.6 | 0.07 | 0.06 | 0.18 |
| **SOL** | 26.0 | 8.5 | 12.1 | 3.8 | 0.07 | **0.03** | 0.56 |
| **TA** | 63.2 | 6.8 | 43.9 | 5.1 | **0.03** | **0.04** | 0.40 |

LG = lateral gastrocnemius; MG = medial gastrocnemius; SOL = soleus; TA = tibialis anterior
CP = cerebral palsy; TD = typically developing
Significant differences ($p<0.05$) are indicated in bold.

### 5.3. Feedback gains

Table S9: Statistical outcome parameters (p-values) for feedback gains for the extended model for children with cerebral palsy and typically developing children.

| **LG** | **p-value** | | |
|---|---|---|---|
| | Level | Group | Level:Group |
| $k_a$ | **0.000** | 0.313 | 0.845 |
| $k_v$ | **0.000** | **0.000** | **0.005** |
| $k_p$ | **0.000** | **0.000** | **0.038** |
| $k'_a$ | **0.037** | **0.045** | 0.912 |
| $k'_v$ | **0.001** | **0.000** | 0.226 |
| $k'_p$ | 0.396 | 0.062 | 0.749 |
| $k_s$ | **0.042** | 0.456 | 0.500 |

| **TA** | **p-value** | | |
|---|---|---|---|
| | Level | Group | Level:Group |
| $k_a$ | **0.000** | **0.023** | 0.119 |
| $k_v$ | **0.000** | **0.000** | **0.012** |
| $k_p$ | 0.067 | 0.247 | 0.092 |
| $k'_a$ | 0.762 | 0.904 | 0.699 |
| $k'_v$ | 0.090 | **0.002** | 0.765 |
| $k'_p$ | 0.150 | **0.022** | 0.840 |

| **MG** | **p-value** | | |
|---|---|---|---|
| | Level | Group | Level:Group |
| $k_a$ | **0.001** | **0.004** | **0.005** |
| $k_v$ | **0.000** | **0.002** | 0.157 |
| $k_p$ | **0.000** | 0.443 | 0.738 |
| $k'_a$ | **0.000** | **0.000** | 0.193 |
| $k'_v$ | **0.001** | **0.000** | 0.212 |
| $k'_p$ | 0.431 | 0.054 | 0.785 |
| $k_s$ | 0.087 | 0.273 | **0.025** |

| **SOL** | **p-value** | | |
|---|---|---|---|
| | Level | Group | Level:Group |
| $k_a$ | **0.000** | **0.018** | 0.326 |
| $k_v$ | **0.000** | **0.016** | 0.435 |
| $k_p$ | **0.003** | **0.020** | 0.271 |
| $k'_a$ | **0.002** | **0.000** | 0.143 |
| $k'_v$ | **0.003** | **0.003** | 0.220 |
| $k'_p$ | 0.919 | 0.198 | 0.978 |
| $k_s$ | 0.099 | 0.219 | 0.127 |

LG = lateral gastrocnemius; MG = medial gastrocnemius; SOL = soleus; TA = tibialis anterior
ka = acceleration gain; kv = velocity gain; kd = displacement gain; ka’ = prime acceleration gain; kv’ = prime velocity gain; kd’ = prime displacement gain; ks = stiction gain.
Significant differences are indicated in bold.

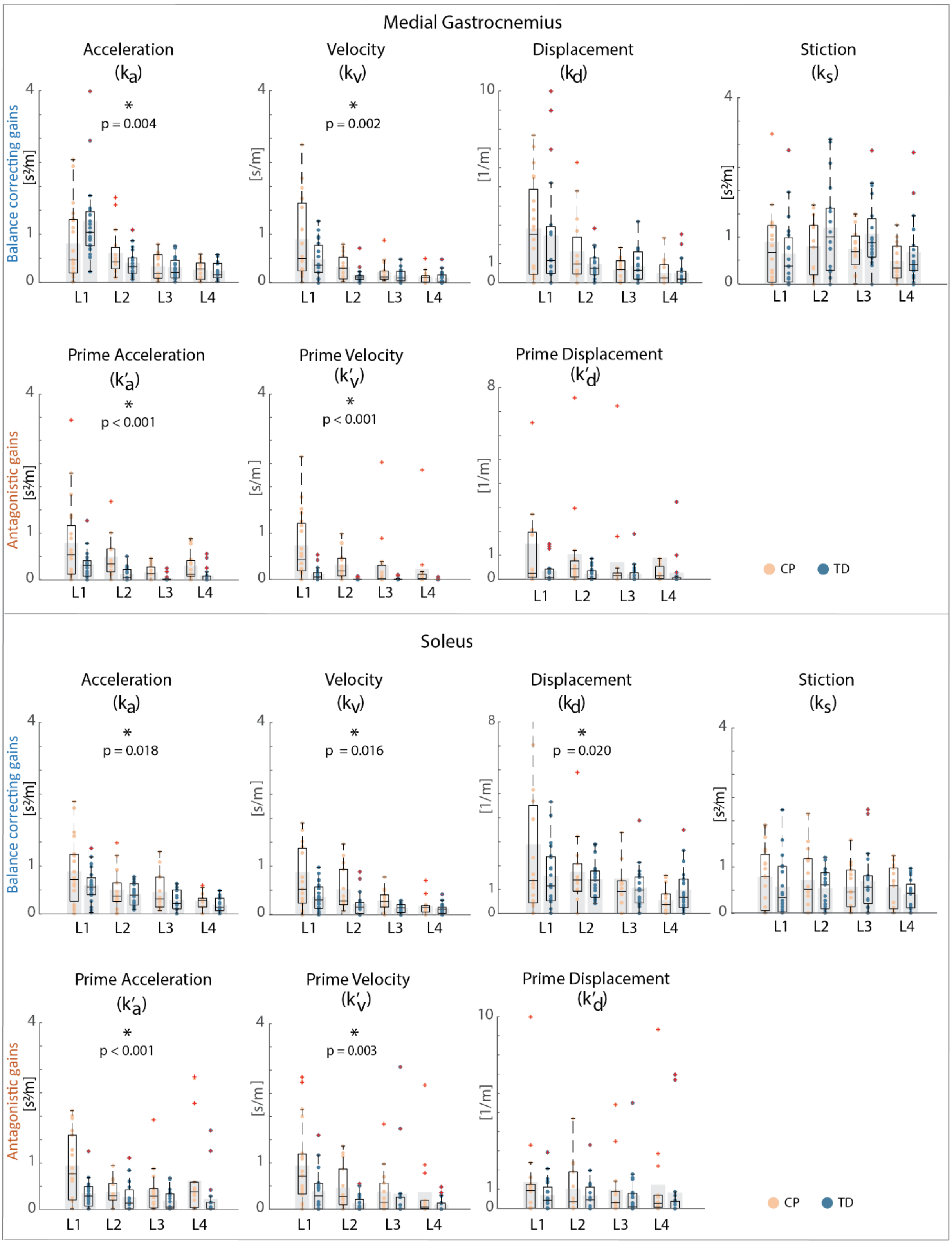


Figure S3: Center of mass feedback gains for all levels for children with cerebral palsy and typically developing children. **a)** Medial gastrocnemius. **b)** Soleus.
Upper row: balance correcting pathway gains, bottom row: antagonistic pathway gains. L1-L4: level 1 to level 4. Grey bars indicate group average, boxplots in black indicate mean and interquartile ranges, and dots represent individual scores. Children with cerebral palsy (CP) in orange, typically developing (TD) children in blue. Significant differences (p< 0.05) between groups are indicated with a star and p-values.

### S6. Correlation with MAS

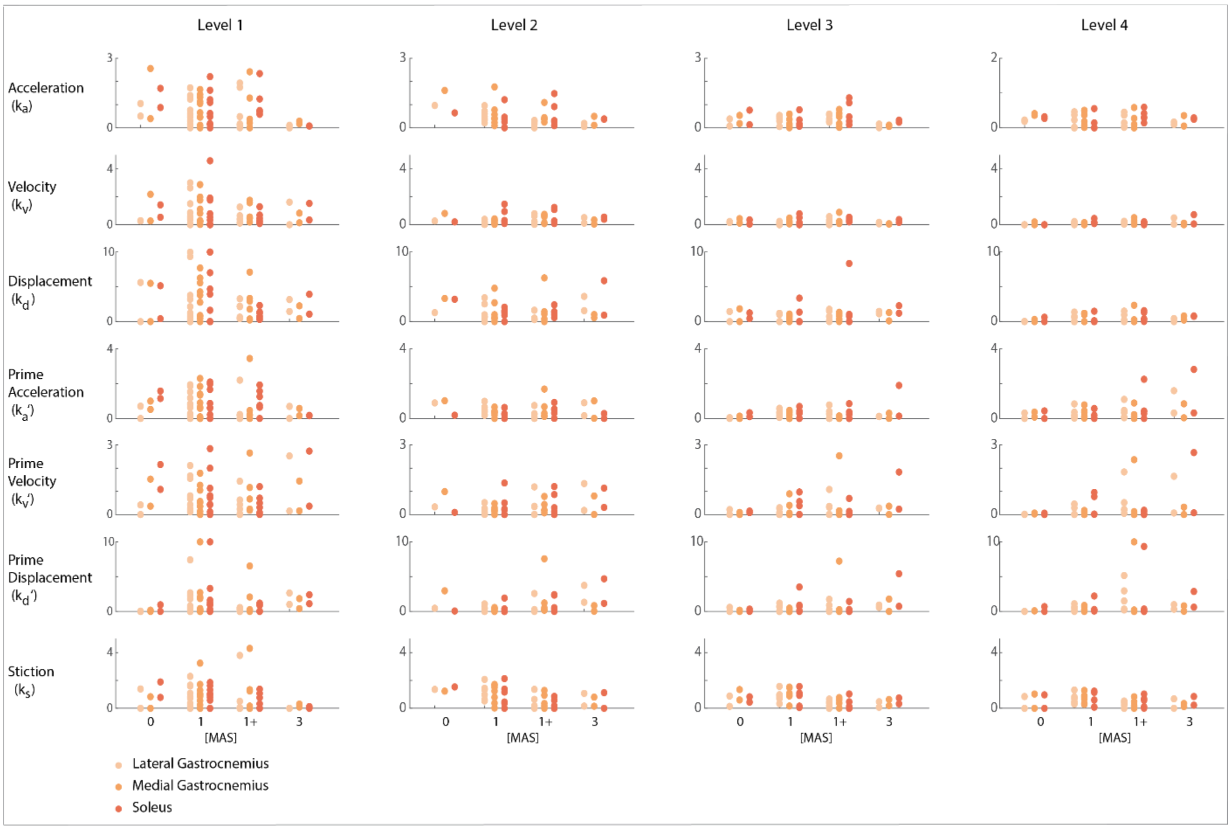


Figure S4: Associations between feedback gains for the lateral gastrocnemius, medial gastrocnemius and soleus and the Modified Ashworth Score of the gastrocnemii. Dots are individual scores. Gains for lateral gastrocnemius in light orange, medial gastrocnemius in orange, soleus in dark orange.

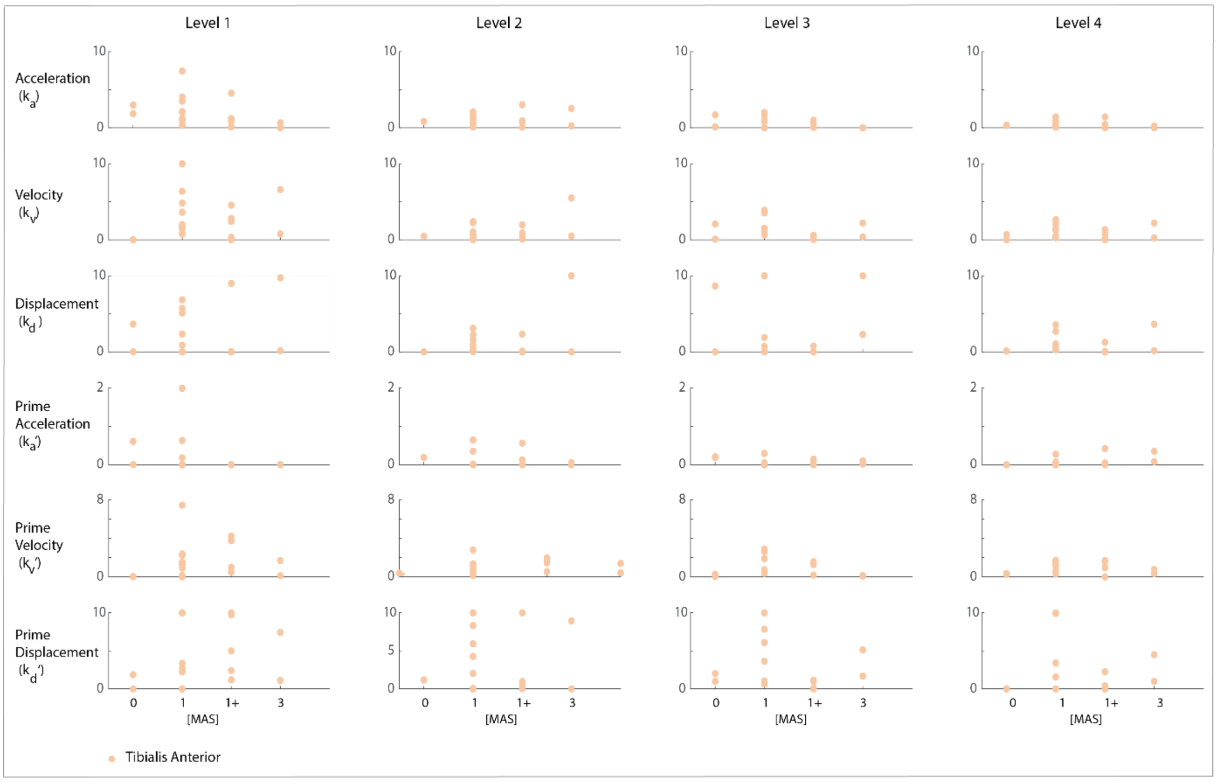


Figure S5: Associations between feedback gains for the tibialis anterior and the Modified Ashworth Score of the gastrocnemii. Dots are individual scores.
